# A CALCIUM-RICH MULTI-MINERAL INTERVENTION TO MODULATE COLONIC MICROBIAL COMMUNITIES AND METABOLOMIC PROFILES IN HUMANS: Results from a 90-day trial

**DOI:** 10.1101/19001255

**Authors:** Muhammad N. Aslam, Christine M. Bassis, Ingrid L. Bergin, Karsten Knuver, Suzanna M. Zick, Ananda Sen, D. Kim Turgeon, James Varani

**Affiliations:** Departments of Pathology, The University of Michigan Medical School; Departments of Internal Medicine, Divisions of Infectious Diseases, The University of Michigan Medical School; The Unit for Laboratory Animal Medicine, The University of Michigan Medical School; Departments of Family Medicine, The University of Michigan Medical School; Department of Nutritional Science, The University of Michigan School of Public Health Ann Arbor, MI 48109; Departments of Internal Medicine, Divisions of Gastroenterology and Biostatistics, The University of Michigan Medical School

**Keywords:** Aquamin, bile acids, colon cancer, colon cancer chemoprevention, calcium, gut microbiome, short chain fatty acids

## Abstract

Aquamin, a calcium-, magnesium-, and multiple trace element-rich natural product has polyp prevention efficacy based on preclinical studies. The overall goal of this study was to determine the safety and tolerability of Aquamin when used as a dietary chemopreventative in humans. Additionally, we determined the effects of Aquamin on the colonic microbial community and attendant metabolomic profile. Thirty healthy male and female human participants were enrolled in a 90-day trial in which the effects of Aquamin (delivering 800 mg of calcium per day) were compared to those of calcium alone or placebo. Before and after the interventional period, colonic biopsies and stool specimens were obtained. All 30 participants completed the study without serious adverse events. There were no changes in liver function markers. Compared to pretreatment values, intervention with Aquamin led to a reduction in total bacterial DNA and a shift in the microbial community. Treatment with calcium alone also produced a decline in total bacteria, but smaller than seen with Aquamin, while no reduction was observed with placebo. In parallel with microbial changes, a reduction in bile acid levels and a slight increase in the level of the short chain fatty acid (SCFA) acetate in stool specimens from Aquamin-treated participants was noted. No change in bile acids or SCFAs was observed with calcium alone or placebo. We conclude from these studies that Aquamin is safe and tolerable in healthy human participants and may produce beneficial alterations in the colonic microbial community and the attendant metabolomic profile.

## INTRODUCTION

Epidemiological studies have shown an inverse relationship between calcium intake and colon cancer incidence (1). Experimental studies in animals have substantiated anti-tumor efficacy in the colon (2) and *in vitro* studies have provided mechanistic insight into how calcium influences epithelial cell proliferation and differentiation (3). In spite of these data, interventional trials with calcium have had only modest success in reducing colon polyp formation, with some trials demonstrating a reduction in incidence (4,5), while others showing essentially no protection (6,7) or an increase in incidence of a more-aggressive sessile-serrated colon polyp phenotype (8). This complicated picture supports the idea that adequate or optimal calcium intake throughout life may be beneficial, but in a realistic dietary setting, the efficacy of supplementation by calcium alone may be limited by dietary complexity. Potentially adverse effects at higher dose levels further complicate the picture (9).

Recent evidence suggests that the anti-neoplastic activity of calcium supplementation may be enhanced by concomitant inclusion of additional trace minerals along with calcium. This has been demonstrated in our own studies using Aquamin, a calcium- and magnesium-rich multi-mineral product obtained from mineralized red marine algae. In two long-term (15-18 month) studies in mice, Aquamin more effectively suppressed colon polyp formation than calcium alone (10,11). Further, Aquamin was more effective than calcium alone at suppressing proliferation and inducing differentiation in human colon carcinoma cells in monolayer culture (12,13), and human adenoma-derived colonoids (14). Thus, Aquamin (as a source of calcium along with magnesium and additional trace minerals) may, ultimately, prove to be a more effective dietary colon polyp chemopreventative than calcium alone.

Exact mechanisms by which calcium alone or multi-mineral supplementation exert anti-neoplastic activity are currently unknown. In addition to direct anti-proliferative and pro-differentiating effects on colonic epithelium, beneficial activity may be mediated indirectly through effects on the gut microbial population and/or changes in gut microbial metabolic activity. The bacterial community in the gastrointestinal tract plays several important roles that contribute to health. A “healthy” microbiome is important for food digestion and production of important metabolites. In addition, a healthy microbial community helps maintain the tissue barrier, regulates the host immune response and provides protection against pathogen overgrowth (15). Dysbiosis, especially in the colon, can lead to barrier breakdown and initiate a chronic inflammatory response. Dysbiosis has been directly linked to inflammatory bowel diseases and indirectly to the formation of premalignant colon polyps (15-17). In previous studies in mice, dietary calcium supplementation has been shown to cause a shift in the gut microbial community in comparison to control (18,19). In another study, Aquamin itself induced gut microbial changes (20).

Metabolic changes may also be important. At high concentrations, bile acids are membrane-active agents and can be cytotoxic (21). In addition, certain bacterially-derived secondary bile acids have carcinogenic properties (22). Alterations in the gut microbial population can affect bile acid profiles (23). In our own murine study (19), calcium supplementation altered the gut microbiome and reduced the level of total bile acids and certain microbially-derived secondary bile acids. In addition to bile acids, gut microbes also produce short chain fatty acids (SCFAs) (e.g. acetate, propionate and butyrate) in the colon. SCFAs have demonstrable protective effect against colonic inflammation and carcinogenesis (24). Thus, a shift in the gut microbial community may affect colon health through alterations in bacterially-derived metabolites.

Whether similar microbial/metabolomic changes might also be seen in human participants is not known. To test the feasibility of using dietary Aquamin as an interventional strategy in humans, we conducted a 90-day, FDA-approved pilot study, comparing daily supplementation with Aquamin to calcium alone and placebo in healthy human participants. While the primary purpose of this study was to assess Aquamin’s tolerability and safety, we also evaluated microbial community structure and assessed bile acid, SFCA and eicosanoid profiles at endpoint in comparison to baseline values in participants from all three groups. Elucidating the microbial and metabolic changes resulting from intervention is important for identifying potential mechanisms of action with Aquamin. Identifying measurable alterations in microbial and metabolomic parameters and determining effect-size with intervention will also be helpful for determining the necessary sample size going forward in a larger-scale clinical trial. Initial results from the current study are reported herein.

## MATERIALS AND METHODS

### Aquamin and control interventions

Aquamin is a calcium- and magnesium-rich natural product obtained from the skeletal remains of red marine algae of the *Lithothamnion* genus. In addition to calcium and magnesium, Aquamin contains detectable levels of 72 additional trace minerals including trace elements from the lanthanide family (essentially all of the minerals accumulated by the algae from seawater). Aquamin is sold as a food supplement (GRAS 000028) (Marigot Ltd, Cork, Ireland) and is used in various products for human consumption in Europe, Asia, Australia, and North America. A single batch of Aquamin-Food Grade^®^ was used for this study. Mineral composition was established via an independent laboratory (Advanced Laboratories; Salt Lake City, Utah). Supplement Table 1 provides a complete list of elements detected in Aquamin and their relative amounts. Calcium carbonate was used for comparison and maltodextrin was used as placebo.

### Study participants

This study was a pilot, double-blind, parallel assignment, randomized clinical interventional trial in which thirty participants were included. Participants were male or non-pregnant female, in general good health, but having “an increased risk for colon cancer” based on i) a personal history of colorectal polyp, early stage (stage I or II) colon cancer treated by surgical removal without recommendation for adjuvant therapy, or stage III colon cancer treated with surgery > 5 years prior or ii) a first degree relative diagnosed under the age of 60 with colorectal cancer. Exclusion criteria included history of kidney disease or kidney stones, Crohn’s disease or ulcerative colitis, gastrointestinal hemorrhagic disorders, or coagulopathy, hereditary non-polyposis coli or familial adenomatous polyposis. The participants were recruited through the Michigan Medicine web portal and by posting flyers in the hospital.

This clinical interventional trial was conducted with FDA approval of Aquamin as an Investigational New Drug (IND#118194) and with oversight by the Institutional Review Board at the University of Michigan Medical School (IRBMED)-IRB#HUM00076276. The study was registered as an interventional clinical trial with details at Clinicaltrials.gov (study identifier NCT02647671). All participants provided written informed consent prior to inclusion. This phase I trial involving human participants was carried out in accordance with recognized ethical guidelines, for example, Declaration of Helsinki, International Ethical Guidelines for Biomedical Research Involving Human Subjects (CIOMS), the Belmont Report and the U.S. Common Rule.

### Study design

Supplement Figure 1 summarizes the study design of this trial in a flowchart. Briefly, at screening participants were given the NIH Diet History Questionnaire II (DHQ II) a food frequency questionnaire which includes portion size and dietary supplement questions as a way to evaluate baseline calcium levels in the past year (25). Participants were also asked about use of dietary supplements, antibiotics and non-steroidal anti-inflammatory drugs. No participants were on antibiotics at or in close proximity to the time of screening. Individuals ingesting supplements containing calcium and / or vitamin D were required to undergo a two-week “wash out” period prior to starting and to not use these supplements during the study participation.

Thirty participants underwent baseline flexible sigmoidoscopy (unprepped; *i.e.*, without bowel cleansing procedure). Twelve 2.5 mm colonic biopsies were obtained along with two stool specimens from within the sigmoid colon (20 cm above the anus). Tissue and stool samples were saved in 10% formalin, cryopreserved for cultures or snap frozen in liquid nitrogen and saved at −80°C. Blood was drawn for liver function / liver injury markers (total albumin, bilirubin, aspartate aminotransferase (AST), alanine aminotransferase (ALT) and alkaline phosphatase (ALKP).

After baseline sigmoidoscopy, participants were randomized to one of three groups. Ten participants were treated daily for 90 days with Aquamin providing 800 mg of calcium per day. Ten participants received 800 mg of calcium carbonate daily and ten participants received maltodextrin as placebo. During the interventional period, participants were contacted by study coordinators on a monthly basis to assess study progress / adherence to the study protocol and to identify unwanted side effects. Compliance was assessed by capsule log entries and by counting unused capsules returned at the end of the study.

At the end of the 90-day intervention period (90±5 days), participants again underwent unprepped flexible sigmoidoscopy and eight colonic biopsies along with two stool specimens were collected and stored as at baseline. Blood was also taken for the same serum markers as at baseline. For each of the two time-points, one biopsy and one stool specimen from each participant was utilized for microbiome analysis and one biopsy and one stool specimen was utilized for metabolomic analysis. The remaining tissue samples were retained for backup, or were used for histological, immunohistochemical and proteomic analyses. The results of the latter analyses will be reported separately.

### Microbial analysis

DNA was isolated from colon and stool samples using the Qiagen MagAttract PowerMicrobiome DNA/RNA kit. Total bacterial DNA levels were estimated using qPCR with broad-range primers BSF8 (AGAGTTTGATCCTGGCTCAG) and BSR357 (CTGCTGCCTYCCGTA), targeting bacterial 16S rRNA genes (26). Each 10 μl qPCR reaction contained 5 μl PowerUp SYBR Green Master Mix (Applied Biosystems by Thermo Fisher Scientific), 4 μl sample DNA (1:40 dilution) and 0.4 μM of primers. The following cycles were run on a Light Cycler 96 (Roche): 1x(2 min at 50°C, 10 min at 95°C), 40x(15 s at 95°C, 1 min at 60°C). Each sample was run in duplicate.

Microbial community profiles were generated by Illumina^®^ MiSeq sequencing of the V4 region of 16S rRNA-encoding genes after amplifying the extracted DNA (27). Samples were amplified, normalized and sequenced on the MiSeq and analysis was performed using the MiSeq SOP (28) for the software mothur (v.1.39.0 and v.1.39.5) as described in our earlier study (19). In case of low bacterial biomass, 3μl of DNA were amplified by touchdown PCR method [1×(2 min at 95°C), 20x(20 s at 95°C, 15 s at annealing temperature (starts at 60°C, decreases 0.3°C/cycle), 5 min at 72°C), 20x(20 s at 95°C, 15 s at 55°C, 5 min at 72°C), 1x(10min at 72°C)]. Thirteen samples, including 8/10 post-interventional Aquamin colon samples, required touchdown PCR (with 40 amplification cycles versus standard PCR with 30 cycles) to achieve sufficient sample amplification for sequencing. After processing, sequences were binned into operational taxonomic units (OTUs) based on 3% difference in sequence using the OptiClust method (29). Comparisons between groups included differences in community structure using OTU-based Yue and Clayton distance metric (θ_YC_) (30). θ_YC_ distances were visualized using principal coordinates analysis (PCoA). Specific OTUs driving community differences were identified by linear discriminant analysis effect size (LEfSe) (31), as was done in our previous study (19). LEfSe utilizes both statistical significance and effect size (linear discriminant analysis score, or LDA) to determine the features (OTUs, in this case) that are differentially abundant between groups (i.e., pre-post differences). Alpha diversity was assessed by change in the Shannon diversity index from baseline. To evaluate Aquamin-associated microbial community differences in major gut phyla, we sorted the 1000 most abundant OTUs in each of the baseline and final visit samples into phyla and compared inter-group differences in each of these phyla at endpoint. The DNA isolation and 16S rRNA gene sequencing were done by the University of Michigan Microbial Systems Molecular Biology Laboratory.

### Metabolomic analysis

Bile acids, SCFAs and eicosanoid composition were quantified in colon biopsies and stool specimens from a randomly chosen subset of participants (n=6 per group) at each time point (pre- and post-intervention). Bile acids were quantified using Liquid Chromatography-Mass spectrometry (LC-MS) in a two-step solvent extraction (32) as performed in the Regional Comprehensive Metabolomics Resource Cores (RCMRC) at the University of Michigan. Supernatants were combined, dried, and re-suspended for LC-MS separation by reverse-phase liquid chromatography (RPLC) and measured by multiple reaction monitoring (MRM). Sample identification was performed by comparison of retention times and mass to an in-house library of bile acids. SCFA analysis was performed by the RCMRC by electron ionization-gas chromatography - mass spectrometry (EI-GCMS) as described recently (33). Compound identification was performed against a library of known SCFAs. Colon tissue was also utilized to analyze a profile of eicosanoids. Eicosanoids were extracted and concentrated using solid phase extraction in colon biopsies. The eluent was dried and re-suspended for LC-MS separation by RPLC and measurements by MRM methods (34). Stool specimens were not analyzed for eicosanoids since these metabolites are not bacterial products. All analytes were reported as pmol/mg, after normalization to the sample weight.

### Statistical analysis

Pre (baseline) vs post (endpoint) comparisons were conducted for each microbial or metabolomic endpoint for individual participants in each of the three treatment groups (placebo, calcium, Aquamin). Group means and standard deviations were then calculated for baseline and endpoint measures. Intergroup differences for normally distributed continuous data were made by ANOVA followed by pairwise group comparisons with Bonferroni corrections for multiple comparisons. Microbial community distances were compared by AMOVA (analysis of molecular variance) within the program mothur (v.1.39.0 and v.1.39.5) (35). For continuous data that failed normality testing intergroup comparisons were made by Kruskal-Wallis ANOVA followed by Mann Whitney-Wilcoxon pairwise comparison. Statistical analyses were performed using GraphPad Prism (version8) and R computing software (R version 3.5.3 and RStudio Version 1.1.463). Due to a small cohort size, analyses were not adjusted to any baseline sociodemographic or clinical characteristics or dietary calcium intake.

## RESULTS

### Participant characteristics

Thirty-six participants were randomly enrolled. Six subjects were lost to follow-up prior to intervention and a total of 30 participants (10 per arm) completed the study. This included of 22 female and 8 male participants. Participants were randomized to study arms without regard to age or gender (placebo: 4 males and 6 females; calcium: 2 males and 8 females; Aquamin: 2 males and 8 females). Ages ranged from 20-66 years. Compliance (capsule intake) was 96% across the three groups. Complete Demographic characteristics are presented in Supplement Table 2. Based on responses to DHQ II, the average dietary calcium intake (mg/day) values for the three groups at baseline were estimated to be: placebo = 817±245; calcium = 964±412 and Aquamin = 919±545. There were no statistical differences in calcium intake among the groups.

### Safety and tolerability

Self-reported adverse events over the course of study are shown in Supplement Table 3. All adverse events were minor (*i.e*., headache, gastrointestinal symptoms) and did not preclude any individual from completing the study. The number of individuals reporting adverse events in the Aquamin group was the same as in the placebo group (3 of 10) while 6 of 10 individuals in the calcium group reported one or more events. No serious adverse events (defined as necessitating cessation of study participation or medical intervention / hospitalization) occurred. Serum liver function / liver injury markers are presented in Supplement Table 4. No significant change in any individual marker was observed over the course of the study. Likewise, there was no difference among the treatment groups in serum calcium levels before and after intervention (Supplement Table 4).

### Effects on bacterial DNA in colon biopsy and stool specimens

qPCR was used to assess bacterial 16S rRNA gene levels in colon biopsy and stool specimens as a way to estimate total bacterial DNA. Figure 1 demonstrates that in both colon biopsy and stool specimens, post-treatment Aquamin samples had higher cycle quantification (Cq) values, indicating lower amounts of total bacterial DNA than at baseline. There was also a decrease in total bacterial DNA in specimens from calcium-treated participants, although, on average, the decrease was not as great as that seen with Aquamin. Placebo specimens showed no (average) decline in DNA content at endpoint.

**Figure 1.**
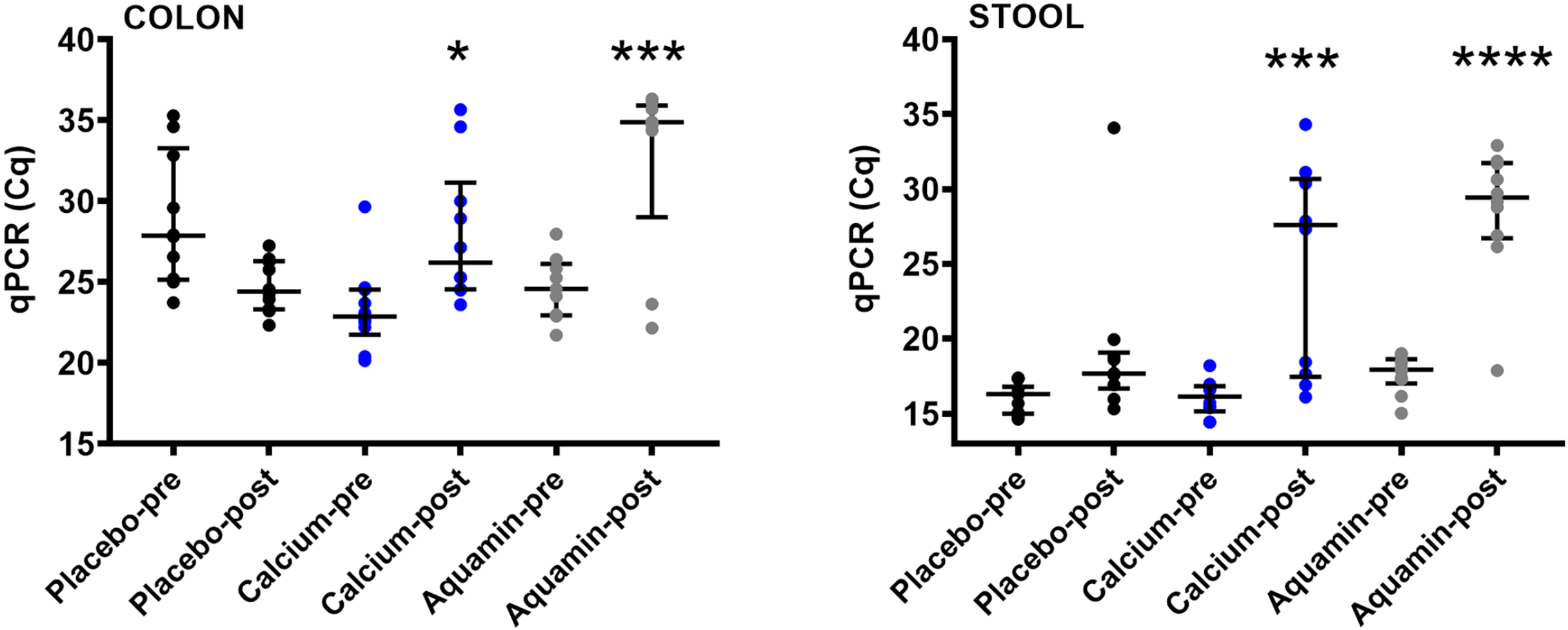
Decrease in bacterial DNA with Aquamin. Quantitative PCR (qPCR) for bacterial DNA in colon (left) and stool (right) samples. The average Cq value from duplicate qPCR runs was plotted except for one post-Aquamin colon sample which failed to reach the amplification threshold in one duplicate and was therefore represented by a single Cq value (34.4) rather than the average. Another post-Aquamin colon sample failed to reach the amplification threshold in both duplicates and was, therefore, not plotted or included in the statistical analysis. A decrease in the total amount of bacterial DNA was indicated by a higher cycle quantification (Cq) value (longer time to reach amplification threshold). There was no difference in pre- vs post-supplementation for the placebo group. * reflects significance at *p*<0.05, *** reflects significance at *p*<0.001 and **** reflects significance at *p*<0.0001 compared to the corresponding pretreatment value.

### Alterations in gut microbial communities

To determine if Aquamin altered the composition of the gut microbiota in the participants, we analyzed bacterial 16S rRNA gene sequences from colon and stool specimens to explore pre-post-interventional differences. After sequence processing and exclusion of 2 samples for low sequence counts, a total of 7,081,672 sequences from 118 samples (median: 59233 ± 23618 (SD) sequences/sample; range 2338 - 114235 sequences/sample) were included in this analysis. Shifts in gut microbial community composition were assessed by calculating θ_YC_ distances between the pre- and post-supplementation communities for each individual. Individual pre-post-distance values, grouped by intervention, are shown in Figure 2. Colonic microbial communities demonstrated a bigger shift in the Aquamin-supplemented group than was observed for either the placebo or calcium-supplemented group. The difference between Aquamin and calcium in the colon biopsy specimens reached the level of statistical significance (p=0.0087). Both figure insets show that the majority of the Aquamin samples (8 of 10 in colon and 7 of 10 in stool) were above the median value for all samples.

**Figure 2.**
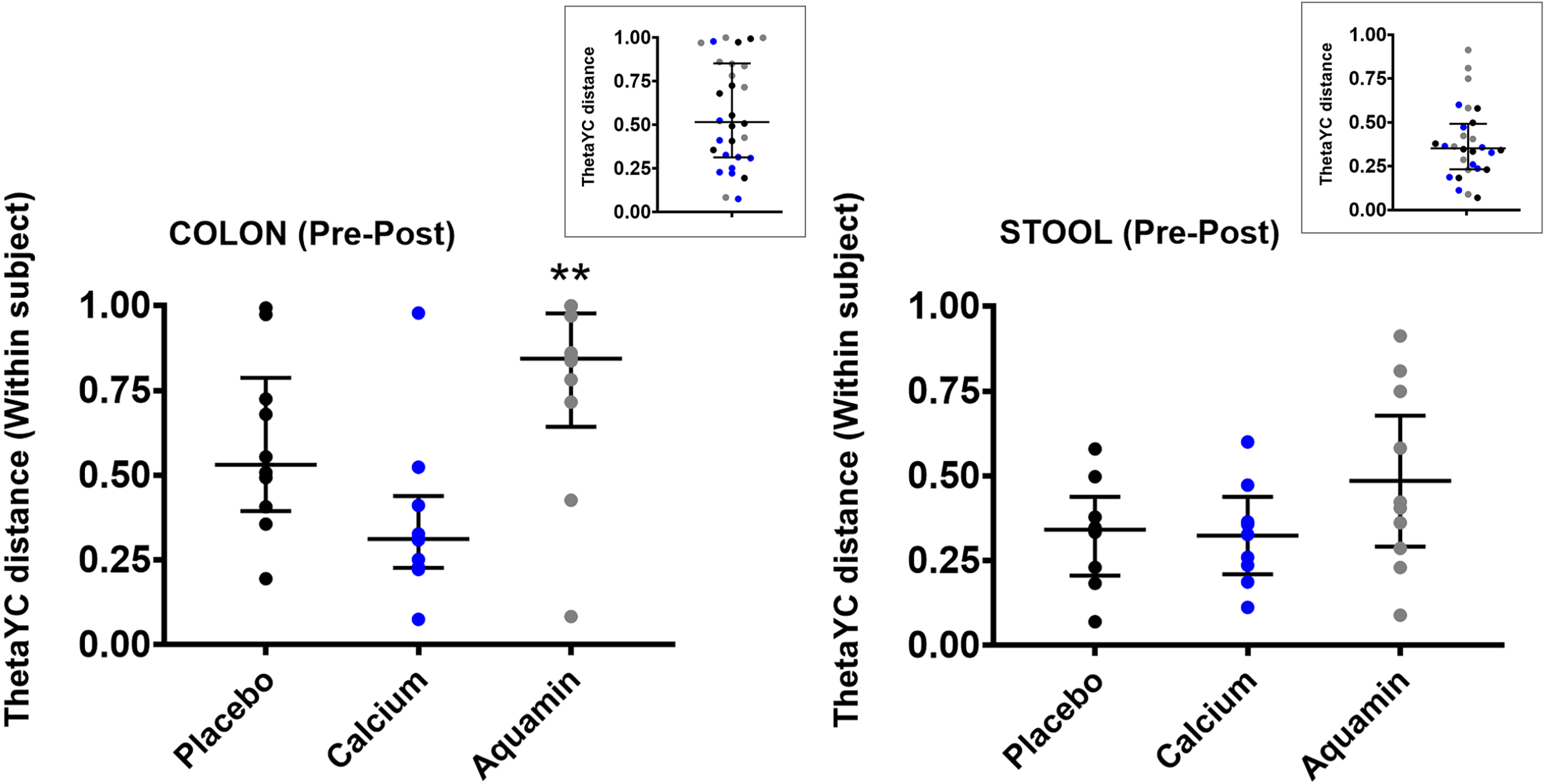
Differences in gut bacterial community composition with Aquamin. θ_YC_ distances as a comparison of pre- vs post-supplementation values in gut bacterial community composition for colon (left) and stool (right) samples. Higher values reflect greater differences. ** reflects significance at *p*<0.01 relative to calcium. The insets show that the majority of the Aquamin (colon and stool) samples were above the median value for all samples. The majority of placebo and calcium samples were at or below the combined median value.

Figure 3 presents principle coordinates analysis (PCoA) data for all three treatment groups in both colon and stool specimens. At baseline, there were no significant differences in the colon bacterial communities (Figure 3A), but following the 90-day intervention period, microbial communities in the colon biopsies from participants treated with Aquamin segregated from those in the placebo or calcium groups (Aquamin versus placebo: AMOVA *p*=0.005 and Aquamin versus calcium: AMOVA *p*=0.009). Further, colonic microbial communities from the post-treatment Aquamin biopsy samples segregated from their own baseline communities (AMOVA *p*=0.022) (Figure 3B), while there was no difference in pre- vs post-supplementation communities for placebo- or calcium-treated participants. In contrast to these results in colon biopsy material, stool sample microbial communities were not different between or within groups either at baseline or at endpoint (Figure 3C and 3D).

**Figure 3.**
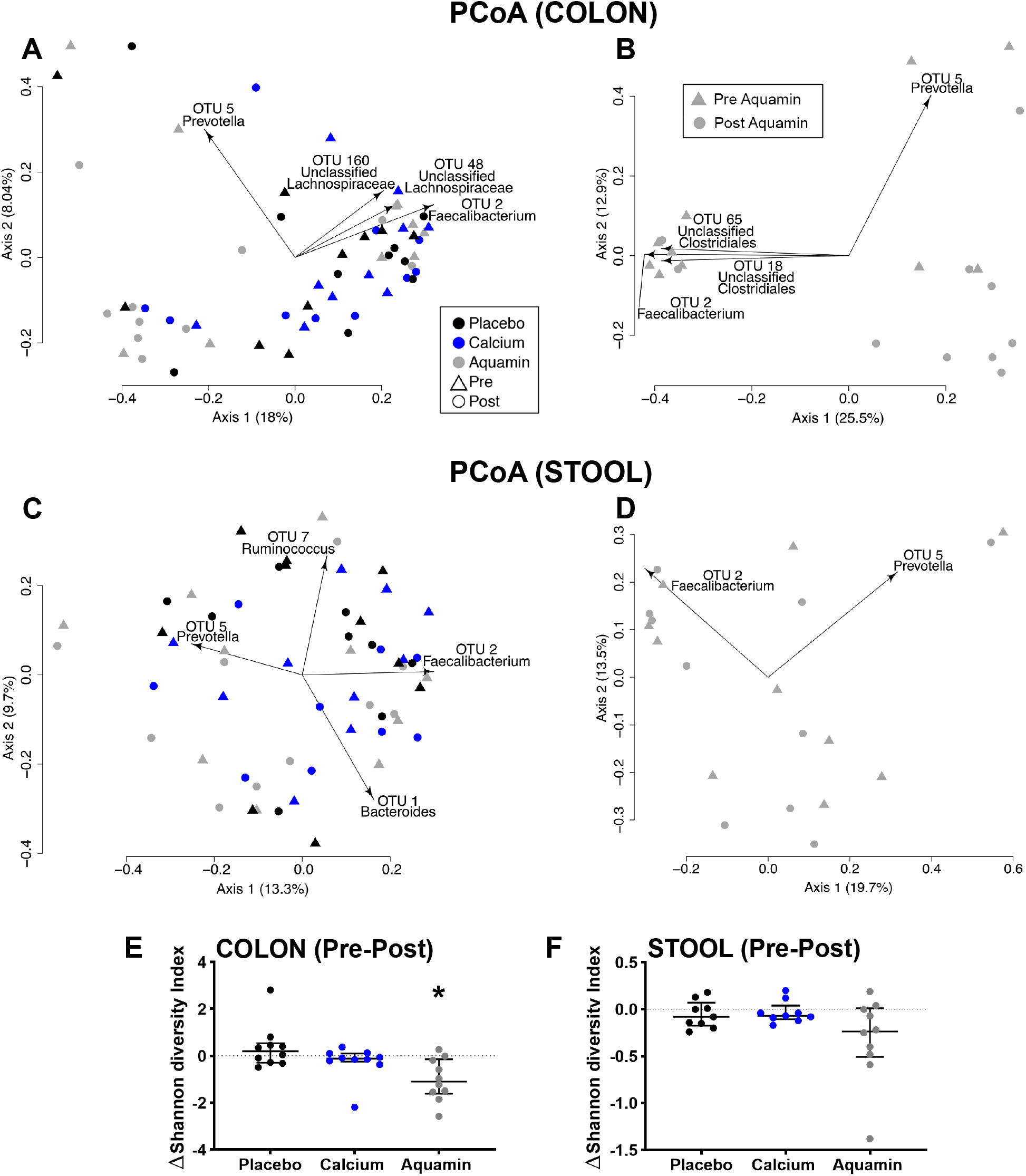
Segregation of gut microbial communities and gut microbial diversity. Biplot figures depicting PCoAs of θ_YC_ distances between colon and stool samples based on Illumina sequencing of the V4 region of 16S rRNA. θ_YC_ Distance is a measure of difference in pre- post-treatment microbial populations from individual participants with each of the three interventions. Data from colon specimens are shown in **A** and **B**, while stool specimen data are presented in **C** and **D**. Some of the OTUs driving the observed segregation between the groups are shown in the biplots. **A**: Significant differences in colon microbiota between post-intervention Aquamin compared to post-intervention placebo and calcium: Aquamin samples with AMOVA *p* values of 0.005 (compared to placebo) and 0.009 (compared to calcium). **B**: Post-intervention colon samples from Aquamin group were also significantly different relative to pre-intervention Aquamin samples with a AMOVA *p* value of 0.022. **C** and **D**. There were no significant differences found in stool samples. **E** and **F**. Shannon diversity index. Aquamin intervention reduced gut microbial diversity as compared to the placebo in colon samples (* reflects significance at *p*<0.05). No significant change was observed in the gut microbial diversity in stool samples.

### Effects on diversity in microbial communities

Alpha diversity was assessed in both colon and stool samples. In Aquamin-treated participants, there was a decrease in diversity (Figure 3E and F). For colon samples, it was significantly low with *p*=0.0037 as compared to the placebo.

### Alterations in the relative abundance of major gut phyla

We assessed alterations in the relative abundance of individual OTUs representing the major gut phyla to compare inter-group differences in each of these phyla at endpoint. *Bacteroidetes, Firmicutes* and *Verrucomicrobia* were decreased with Aquamin while *Actinobacteria* and *Proteobacteria* were increased (Figure 4). Trends were similar in both colon and stool specimens. *Firmicutes* OTUs also showed a drop with calcium intervention (colon biopsies only) but there was little change in the other phyla. Little change in OTU relative abundance in any of the major phyla was seen with placebo.

**Figure 4.**
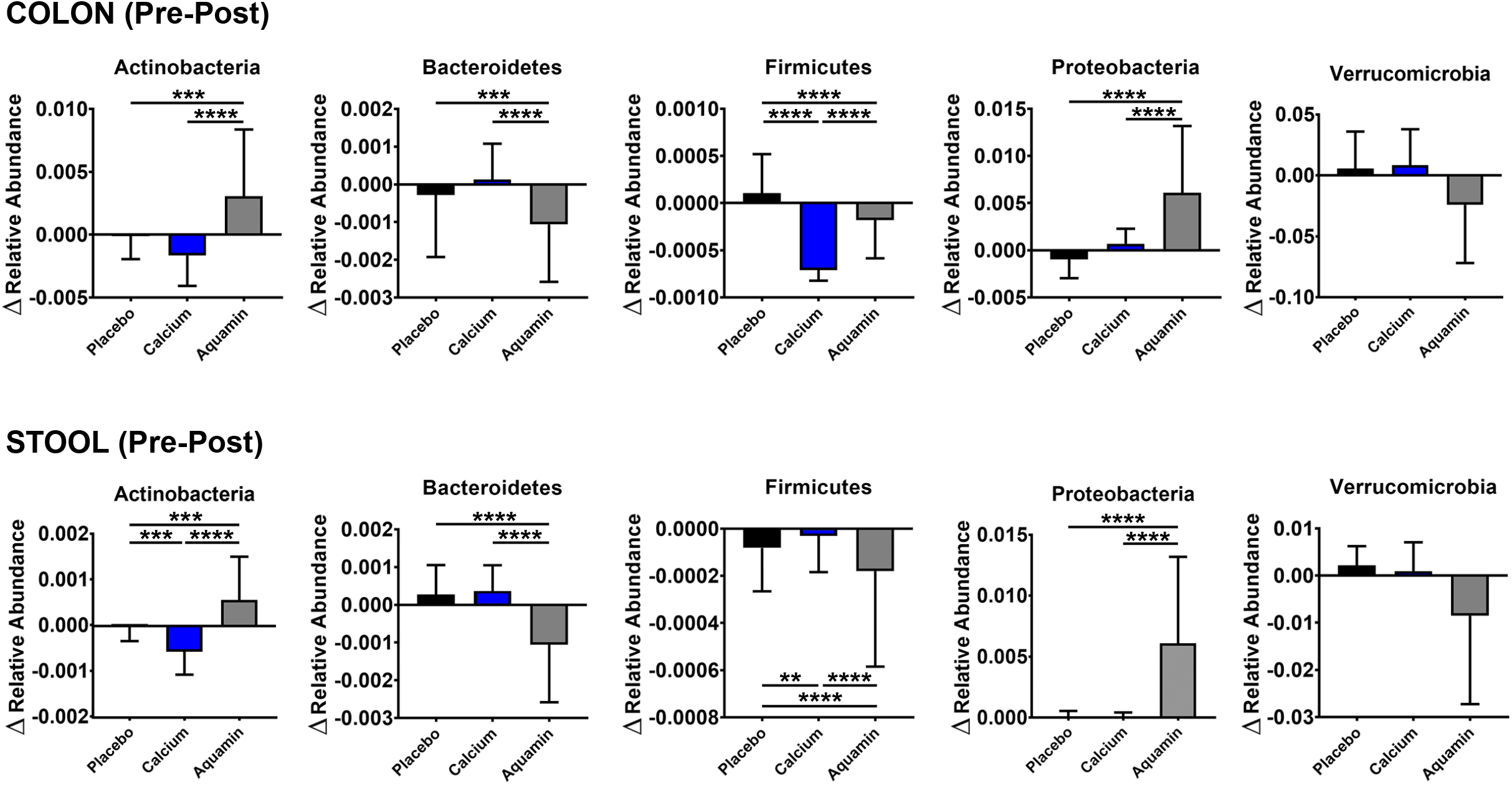
Alterations in the relative abundance of major gut phyla with Aquamin supplementation. The change in the relative abundances of the top 1000 OTUs (pooled by phyla) and assessed by pre-post-intervention analysis among three interventions in colon and stool specimens. For this analysis, 43-48 OTUs across the three treatment groups were pooled for *Actinobacteria*, 101-110 OTUs for *Bacteroidetes*, 543-591 OTUs for *Firmicutes*, 31-39 OTUs for *Proteobacteria* and 4-6 OTUs for *Verrucomicrobia* phyla. ** reflects significance at *p*<0.01, *** reflects significance at *p*<0.001 and **** reflects significance at *p*<0.0001.

### Identification of specific OTUs driving differences

The specific OTUs were identified by LefSe to explain the pre-post-supplementation differences with Aquamin treatment. In the Aquamin supplemented group, 36 OTUs with an LDA score >3 and significance *p*<0.05 were identified in colon samples as shown in Tables 1A and 1B. Overall, more genera decreased than increased after supplementation. Differences between pre- and post-supplementation were driven principally by an increases in OTUs within the normally less abundant phyla *Proteobacteria*, and *Actinobacteria*, along with three OTUs of phylum *Firmicutes*. (Tables 1A) and a decrease in OTUs within the normally higher abundance phyla *Firmicutes* and *Bacteroidetes* (Tables 1B). A complete list of altered OTUs in colon and stool samples with an LDA score >2.0 and significance *p*<0.05 are presented in Supplement Table 5.

**Table 1A.**
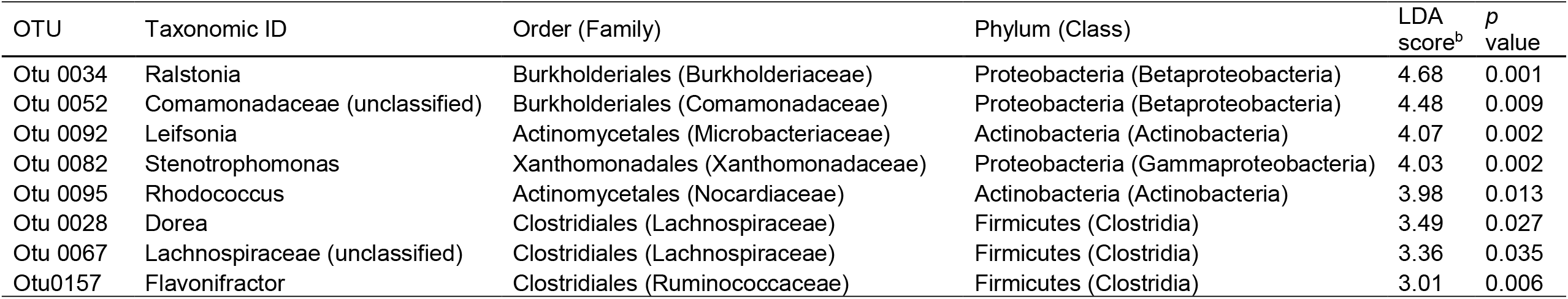
Linear discriminant analysis effect size (LefSe) data for OTU with elevated relative abundance in colon post-Aquamin^a^

| OTU | Taxonomic ID | Order (Family) | Phylum (Class) | LDA score<sup>b</sup> | *p* value |
| --- | --- | --- | --- | --- | --- |
| Otu 0034 | Ralstonia | Burkholderiales (Burkholderiaceae) | Proteobacteria (Betaproteobacteria) | 4.68 | 0.001 |
| Otu 0052 | Comamonadaceae (unclassified) | Burkholderiales (Comamonadaceae) | Proteobacteria (Betaproteobacteria) | 4.48 | 0.009 |
| Otu 0092 | Leifsonia | Actinomycetales (Microbacteriaceae) | Actinobacteria (Actinobacteria) | 4.07 | 0.002 |
| Otu 0082 | Stenotrophomonas | Xanthomonadales (Xanthomonadaceae) | Proteobacteria (Gammaproteobacteria) | 4.03 | 0.002 |
| Otu 0095 | Rhodococcus | Actinomycetales (Nocardiaceae) | Actinobacteria (Actinobacteria) | 3.98 | 0.013 |
| Otu 0028 | Dorea | Clostridiales (Lachnospiraceae) | Firmicutes (Clostridia) | 3.49 | 0.027 |
| Otu 0067 | Lachnospiraceae (unclassified) | Clostridiales (Lachnospiraceae) | Firmicutes (Clostridia) | 3.36 | 0.035 |
| Otu0157 | Flavonifractor | Clostridiales (Ruminococcaceae) | Firmicutes (Clostridia) | 3.01 | 0.006 |

**Table 1B.** Linear discriminant analysis effect size (LefSe) data for OTU with decreased relative abundance in colon post-Aquamin^a^

| OTU | Taxonomic ID | Order (Family) | Phylum (Class) | LDA score<sup>b</sup> | *p* value |
| --- | --- | --- | --- | --- | --- |
| Otu 0001 | Bacteroides | Bacteroidales (Bacteroidaceae) | Bacteroidetes (Bacteroidia) | 4.40 | 0.016 |
| Otu 0004 | Blautia | Clostridiales (Lachnospiraceae) | Firmicutes (Clostridia) | 4.15 | 0.023 |
| Otu 0008 | Fusicatenibacter | Clostridiales (Lachnospiraceae) | Firmicutes (Clostridia) | 4.00 | 0.005 |
| Otu 0009 | Anaerostipes | Clostridiales (Lachnospiraceae) | Firmicutes (Clostridia) | 3.92 | 0.032 |
| Otu 0031 | Akkermansia | Verrucomicrobiales (Verrucomicrobiaceae) | Verrucomicrobia (Verrucomicrobiae) | 3.88 | 0.021 |
| Otu 0026 | Ruminococcus | Clostridiales (Lachnospiraceae) | Firmicutes (Clostridia) | 3.83 | 0.008 |
| Otu 0016 | Parabacteroides | Bacteroidales (Porphyromonadaceae) | Bacteroidetes (Bacteroidia) | 3.81 | 0.013 |
| Otu 0043 | Lachnospiraceae (unclassified) | Clostridiales (Lachnospiraceae) | Firmicutes (Clostridia) | 3.80 | 0.027 |
| Otu 0029 | Bacteroides | Bacteroidales (Bacteroidaceae) | Bacteroidetes (Bacteroidia) | 3.68 | 0.006 |
| Otu 0020 | Bacteroides | Bacteroidales (Bacteroidaceae) | Bacteroidetes (Bacteroidia) | 3.63 | 0.029 |
| Otu 0018 | Clostridiales (unclassified) | Clostridiales (Clostridiales_unclassified) | Firmicutes (Clostridia) | 3.60 | 0.024 |
| Otu 0076 | Lachnospiraceae (unclassified) | Clostridiales (Lachnospiraceae) | Firmicutes (Clostridia) | 3.52 | 0.003 |
| Otu 0019 | Blautia | Clostridiales (Lachnospiraceae) | Firmicutes (Clostridia) | 3.51 | 0.004 |
| Otu0010 | Collinsella | Coriobacteriales (Coriobacteriaceae) | Actinobacteria (Actinobacteria) | 3.33 | 0.048 |
| Otu0070 | Lachnospiraceae_unclassified | Clostridiales (Lachnospiraceae) | Firmicutes (Clostridia) | 3.29 | 0.001 |
| Otu0045 | Coprococcus | Clostridiales (Lachnospiraceae) | Firmicutes (Clostridia) | 3.27 | 0.009 |
| Otu0025 | Streptococcus | Lactobacillales (Streptococcaceae) | Firmicutes (Bacilli) | 3.18 | 0.018 |
| Otu0110 | Parasutterella | Burkholderiales (Sutterellaceae) | Proteobacteria (Betaproteobacteria) | 3.16 | 0.021 |
| Otu0054 | Ruminococcus2 | Clostridiales (Lachnospiraceae) | Firmicutes (Clostridia) | 3.15 | 0.031 |
| Otu0107 | Lachnospiraceae_unclassified | Clostridiales (Lachnospiraceae) | Firmicutes (Clostridia) | 3.14 | 0.002 |
| Otu0087 | Bacteroides | Bacteroidales (Bacteroidaceae) | Bacteroidetes (Bacteroidia) | 3.08 | 0.019 |
| Otu0037 | Dialister | Selenomonadales (Veillonellaceae) | Firmicutes (Negativicutes) | 3.08 | 0.019 |
| Otu0069 | Coprococcus | Clostridiales (Lachnospiraceae) | Firmicutes (Clostridia) | 3.07 | 0.045 |
| Otu0074 | Bacteroides | Bacteroidales (Bacteroidaceae) | Bacteroidetes (Bacteroidia) | 3.06 | 0.007 |
| Otu0068 | Lachnospiraceae_unclassified | Clostridiales (Lachnospiraceae) | Firmicutes (Clostridia) | 3.03 | 0.009 |
| Otu0091 | Lachnospiraceae_unclassified | Clostridiales (Lachnospiraceae) | Firmicutes (Clostridia) | 3.03 | 0.027 |
| Otu0128 | Clostridiales_unclassified | Clostridiales (Clostridiales) | Firmicutes (Clostridia) | 3.01 | 0.004 |
| Otu0066 | Alistipes | Bacteroidales (Rikenellaceae) | Bacteroidetes (Bacteroidia) | 3.00 | 0.048 |

<sup>a</sup> Criteria for inclusion: LDA values greater than 3 with $p < 0.05$
<sup>b</sup> LDA values reported as absolute value.
OTU: Operational taxonomic unit

### Effects on bile acid concentrations

Alterations in bile acid levels were assessed in stool samples from Aquamin- and calcium-treated participants in comparison to placebo as shown in Figure 5. Aquamin treatment resulted in a net decline in total bile acids with the major portion of this change being in unconjugated forms (the major components of the fecal bile acid pool). The major unconjugated primary bile acids - *i.e*., cholic acid (CA) and, particularly, chenodeoxycholic acid (CDCA) (measured concurrently with deoxycholic acid [DCA]) - were significantly lower in post-treatment Aquamin stool samples. This was not observed with calcium.

**Figure 5.**
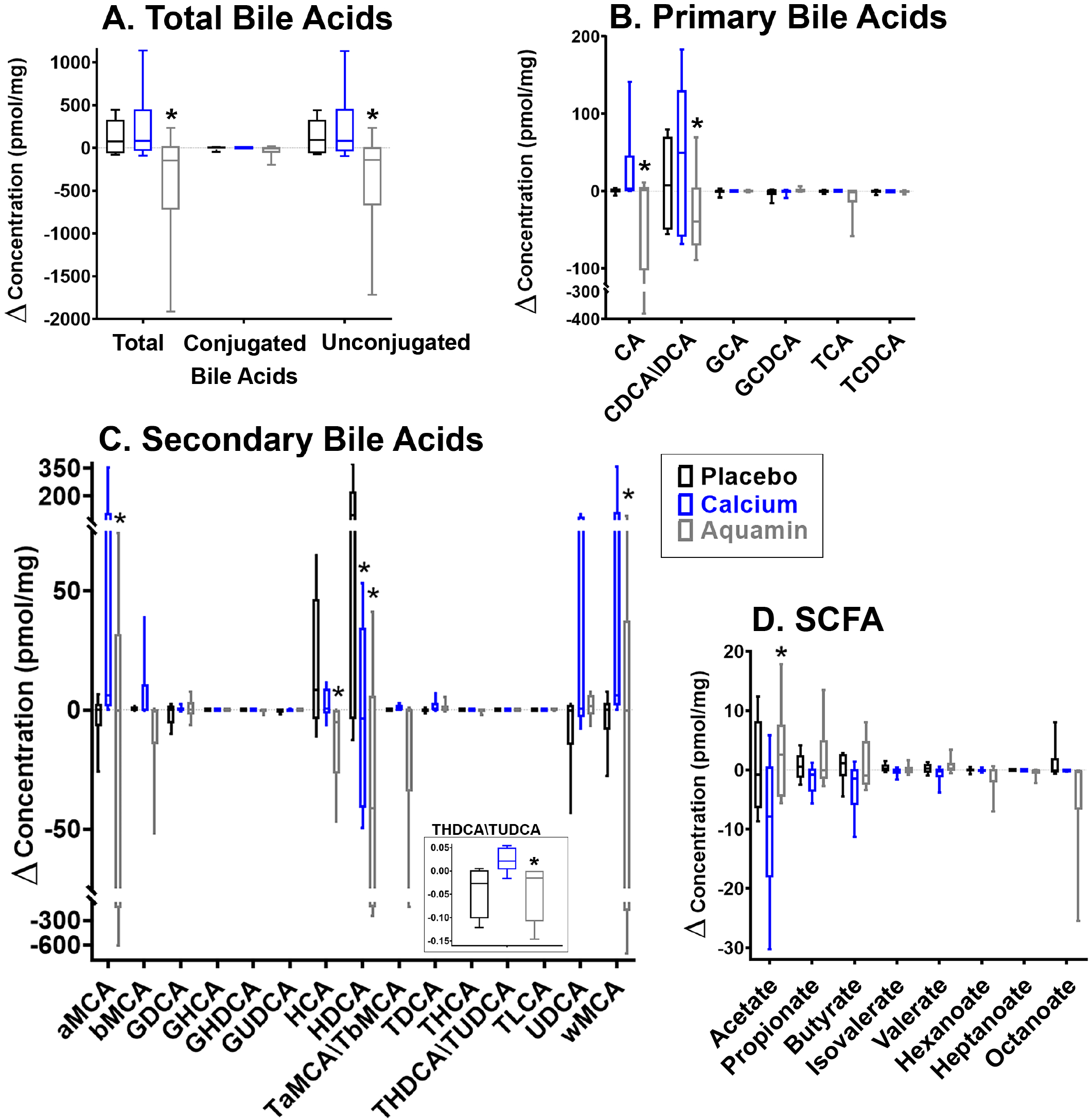
Decrease in Bile acids and increase in SCFAs in stool specimens. Bile acids and SCFA were assessed as described in the Materials and Methods Section. Values shown represent differences in concentration between pre-intervention and post-intervention samples. Asterisks represent statistical significance. **A**: Total bile acids (sum of the total conjugated and total unconjugated bile acids) are shown along with conjugated and unconjugated forms. *reflects decrease with Aquamin at *p*=0.0375 (total) and at *p*=0.0527 (unconjugated) versus calcium. **B**: Primary bile acids. Cholic acid (CA) and chenodeoxycholic acid (CDCA) measured concurrently with deoxycholic acid (DCA) were significantly decreased with Aquamin at *p=*0.0074 (CA) and *p*=0.0310 (DCA/CDCA) versus calcium. **C**: Secondary bile acids. Hyodeoxycholic acid (HDCA) was significantly decreased with Aquamin and calcium versus placebo with *p* value <0.0001 and =0.0149 respectively. Hyocholic acid (HCA) and Taurine-conjugated ursodeoxycholic acid (TUDCA), measured concurrently with taurohyodexycholic acid (THDCA) were decreased with Aquamin relative to calcium, while alpha and omega muricholic acids were also reduced relative to calcium. For HCA, *p*=0.015; for TUDCA/THDCA, *p*=0.013; for *α*MCA, *p=*0.0012 and for ΩMCA, *p*=0.0003. With calcium supplementation, HDCA was also decreased relative to placebo (*p*=0.0149). Inset; TUDCA/THDCA. **D**: SCFA. Acetate was significantly increased with Aquamin relative to calcium alone (*p*<0.0001).

Of the measurable secondary bile acids, taurine-conjugated ursodeoxycholic acid (TUDCA), measured concurrently with taurohyodexycholic acid (THDCA) was decreased, as were the muricholic acids (minor secondary bile acid components). Two other unconjugated secondary bile acids (hyocholic acid [HCA] and hyodeoxycholic acid [HDCA]) were significantly decreased with Aquamin as compared to placebo. HDCA is a byproduct of gut microbial metabolism (36), utilizing HCA and muricholic acids after microbial deconjugation and enzymatic modification. Overall, these Aquamin-associated bile acid changes in stool are consistent with both a decreased bile acid pool and decreased bacterial conversion of primary to secondary bile acids. Of interest, these effects were unique to Aquamin. Calcium supplementation did not show a measurable effect on total fecal bile acids and only HDCA was significantly decreased.

Bile acid levels were also assessed in colon samples from Aquamin- and calcium-treated groups in comparison to placebo (Supplement Figure 2). Similar trends to those seen in stool specimens were observed but none of the changes with Aquamin or calcium intervention reached statistical significance in comparison to placebo except HDCA which was significantly reduced as compared to placebo.

### Effects on SCFA concentrations

SCFA levels were assessed in both stool and colon samples. Apart from a modest (but statistically significant) increase in acetate in stool samples from Aquamin-treated individuals relative to calcium, no other statistically significant alterations were observed in either stool (Figure 5D) or colon (Supplement Figure 2) specimens.

### Effects on colonic eicosanoid concentrations

There was no significant change from baseline detected for any eicosanoid apart from an increase in 13S-hydroxy-octadecadienoic acid (13S-HODE) in calcium-supplemented colon samples (Supplement Figure 3).

## DISCUSSION

This study demonstrated that 90-day dietary intervention with Aquamin was well-tolerated by healthy individuals, and resulted in measurable changes in the colonic microbial community and the attendant bile acid profile. Although the primary purpose of this small phase I trial was safety and tolerability rather than efficacy or mechanisms of action, the identification of Aquamin-associated microbial and metabolomic alterations suggest hypotheses for future studies investigating potential mechanisms of efficacy. The microbial and metabolomic findings should also prove useful as biomarkers going forward in a longer-term interventional trial.

The microbial population present in the colon of an individual is sensitive to environmental influences, especially diet (37,38). Past studies have demonstrated that calcium (18,19) and Aquamin itself (20) can influence the colonic microbial community in mice. In this study, we found that Aquamin supplementation in human participants resulted in a decrease in total colonic microbial DNA and an overall decrease in OTUs within the major gut phyla *Firmicutes* and *Bacteroidetes*, with a decrease in gut microbial diversity. Thus the major impact may simply be a decrease in total gut bacteria. The impact of the decrease in bacteria on colonic health is unknown at this time but poses interesting questions for future exploration. Germ-free mice have shown increased resistance to chemical or dietary-induced colon cancers (39). Since our own previous studies have shown that Aquamin-treated mice have decreased incidence of colon polyps (10,11) as well as fewer liver tumors (40), the possibility that Aquamin-associated microbial decreases may play a mechanistic role in these observations warrants further exploration. Of interest, many of the observations made with samples from Aquamin-treated participants were not seen in samples from individuals ingesting calcium alone. This argues that while calcium is the major mineral component in Aquamin, the effects of Aquamin on the gut microbial community cannot be attributed to calcium alone.

The changes we observed in the post-Aquamin-treatment specimens are similar to the effects of broad-spectrum antibiotic treatment, which have been shown to cause a significant reduction in the proportions of *Firmicutes* and *Bacteroidetes* and an increase in *Proteobacteria* (41). As a follow-up to the observations reported here, it would be informative to assess the potential anti-microbial activity of Aquamin supplementation directly in a minimum inhibitory concentration assay. Metals, particularly cations, have previously been utilized as antimicrobial agents (42). For example, colloidal or nanoparticulate silver and mineral-rich clays are known to have broad-spectrum anti-microbial effects (43,44). Notably, in our study, the administration of Aquamin did not result in diarrhea but there was a drop in bacterial diversity in colon samples; both common adverse effects of broad-spectrum antibiotic use. Thus the anti-microbial effects of Aquamin may be mild in comparison to antibiotics and may allow some of the benefit of reduced gut microbial populations (e.g. reduced potentially carcinogenic secondary bile acids).

In parallel with microbial changes, Aquamin administration decreased the levels of total bile acids and selected primary and secondary bile acids. This finding is consistent with the decrease in total bacteria and may be potentially beneficial to colon health. At high concentrations, bile acids are membrane-damaging and cytotoxic (21). In addition, several secondary bile acids, most notably LCA and DCA (22,23), are known to be carcinogenic. Although these species could not be individually measured in this study for methodological reasons, the combination of DCA and CDCA (the primary bile acid precursor of LCA) was significantly lower in the Aquamin-supplemented group. This suggests that one beneficial effect of Aquamin supplementation might be through effects on bile acid metabolism. A caveat in the interpretation of bile acid data is that it is not currently known whether the reduction in gut bile acid pools is due to altered bacterial composition or, potentially, to a direct effect on the bile acids, themselves. Previous studies have shown that calcium, the major component of Aquamin, has the capacity to precipitate bile acids (45). While mineral binding and precipitation might interfere with detection, this could be beneficial if it prevented potentially harmful secondary bile acids from binding to colonic epithelium or entering the circulation. Arguing against direct binding and precipitation is the finding that calcium alone did not mimic the effects of Aquamin on the total bile acid pool.

A decrease in gut bacteria could, potentially, be harmful to colon health if it resulted in decreased concentration of protective bacterial metabolites, such as the colon-protective SCFAs (24). However, in this study there was no decrease in the colonic concentration of butyrate in the Aquamin-treated group and an actual increase in acetate. Thus, even with the apparent decline in total bacteria, the size and / or composition of the microbial pool was sufficient to maintain pre-supplementation levels of certain SFCAs.

While the data strongly suggest an Aquamin-associated decrease in gut microbial bacteria, the specific shifts in gut microbial populations are harder to interpret. One complication of low microbial biomass samples during microbial sequencing is greater likelihood of detecting reagent or laboratory contaminant microbes in the amplification step. This has been seen in low-biomass samples such as glacier ice, air, rocks, etc (46). While it is not likely to be a major issue in the normal high biomass of the colon, it may be an issue in samples where the biomass has been reduced by antibiotics or other intervention. For this reason, it remains to be seen in further studies whether the elevation in *Proteobacteria* and *Actinobacteria* seen here have relevance as a specific Aquamin-related effect *in vivo* or whether the more important finding is the overall reduction in major gut phyla and decreased overall colonic bacterial biomass.

It should be noted that the antimicrobial effect of Aquamin might not be a direct consequence of the mineral components making up the supplement. Our recent studies employing colonoid culture technology demonstrated up-regulation of proteins having anti-microbial activity upon treatment with Aquamin (47). These included lactotransferrin and natural resistance-associated macrophage protein-1. Lactotransferrin has bacteriostatic activity related to sequestration of free iron and bactericidal properties leading to the release of lipopolysaccharides from the bacterial outer membrane (48). Natural resistance-associated macrophage protein-1 is a divalent transition metal (iron and manganese) transporter involved in iron metabolism. It has been reported to promote host resistance to certain pathogens (49). Thus it is possible that the antimicrobial effects seen with Aquamin *in vivo* are mediated, in part, through its effects on the colonic mucosa, itself. Finally, decreased bacteria, especially as it relates to colon biopsy specimens, might reflect altered bacterial adhesion. In our colonoid culture study (47), numerous proteins involved in cell-cell and cell-matrix adhesive function were altered by Aquamin. Studies by others have noted that alterations in mucosal surface proteins affect microbial interactions with the mucosal wall (15,16).

In addition to the microbial and metabolomic changes, there are other, direct, beneficial effect of Aquamin on the epithelium of the colonic mucosa. Specifically, Aquamin suppressed colon epithelial proliferation and induced differentiation in colonoid culture. Effects on proliferation were seen, primarily, in colonoids derived from large adenomas (14) while improved differentiation – including up-regulation of multiple cell-cell and cell-matrix adhesion molecules and barrier proteins - was observed in normal tissue-derived colonoids (47) as well as in adenoma colonoids (14). Direct effects on the colonic mucosa and indirect effects resulting from changes in microbial / metabolomic profiles are, of course, not mutually exclusive. Further, as noted above, the indirect effects (*i.e*., anti-microbial activity) may reflect changes induced in the colonic mucosa.

Limitations in this study include the small sample size and short duration, which were by design, as this was an initial tolerability study rather than an efficacy study. Although the study was too short and too small to assess efficacy as a colon cancer preventative, it was sufficiently powered to detect significant differences in several microbial and metabolomic (bile acid) endpoints in Aquamin-treated participants relative to the other two interventions. To better define effects on efficacy-related endpoints like inflammatory markers and epithelial differentiation / proliferation indices over a longer period, we are currently beginning a 180-day interventional trial with Aquamin in participants with ulcerative colitis in remission.

In summary, this pilot study demonstrated that 90-day dietary intervention with a calcium- and magnesium-rich multi-mineral supplement was well-tolerated in healthy human volunteers. Adverse events were mild, and largely consisted of gastrointestinal symptoms (constipation, diarrhea etc.) that did not preclude completion of the study. Reports of adverse events were actually less frequent with Aquamin than with calcium alone. Consistent with this, liver function / liver injury markers did not vary significantly with intervention. With respect to microbial and metabolomic findings, Aquamin supplementation resulted in an overall decrease in gut microbial numbers and a decrease in bile acids, including potentially carcinogenic secondary bile acids or their precursors. Despite the anti-microbial-like effects, concentrations of the colon-protective SCFAs were maintained. These findings, along with previous *in vitro* and animal data (10-14,47) showing a beneficial effect of Aquamin on colonic health, support future longer-term interventional studies in human participants. Finally, the observation that Aquamin had a more pronounced effect on gut microbial populations and bile acid levels than calcium alone supports the view that the beneficial activity of Aquamin (calcium in conjunction with additional trace elements) cannot be attributed to calcium alone. This conclusion supports findings from a recent epidemiological study suggesting that calcium in combination of additional minerals may be linked with the lower risk of colorectal cancer (50).

## Data Availability

Microbial sequencing data will be available from the NCBI Sequence Read Archive database, accession number is pending.
Raw bile acid (metabolomic) data will be available in UCSD Metabolomics Workbench, a metabolomic repository, study ID is pending.

## Acknowledgments

This study was supported by NIH grants CA201782 to JV, UL1TR002240 to the Michigan Institute for Clinical & Health Research and 2 P30 DK089503-06 to the Michigan Nutrition and Obesity Research Center (MNORC). Metabolomic work was supported by Pilot/Feasibility Grant from MNORC to IB and MNA. This study also used services at University of Michigan which were supported by NIH funding (UL1TR002240; KL2TR002241; and TL1TR002242) to the Michigan Institute for Clinical & Health Research (MICHR). We thank MICHR, the Michigan Clinical Research Unit (MCRU), the University of Michigan Microbial Systems Molecular Biology Laboratory and the Regional Comprehensive Metabolomics Resource Cores (RCMRC) at the University of Michigan for help with various aspects of the study. We thank our study coordinators (Elaine Brady and Deepa Chandhrasekhar) for their assistance with the study. We also thank Marigot LTD (Cork, Ireland) for providing Aquamin^®^ as a gift.

## Bile acid abbreviations

aMCA: *alpha*-Murocholate
bMCA: *beta*-Muricholate
CA: Cholate
CDCA: Chenodeoxycholate
DCA: Deoxycholate
GCA: Glycocholate
GCDCA: Glycochenodeoxycholate
GDCA: Glycodeoxycholate
GHCA: Glycohyocholate
GHDCA: Glycohyodeoxycholate
GUDCA: Glycoursodeoxycholate
HCA: Hyocholate
HDCA: Hyodeoxycholate
LCA: Lithocholate
TaMCA: Tauro-*alpha*-muricholate
TbMCA: Tauro-*beta*-muricholate
TCA: Taurocholate
TCDCA: Taurochenodeoxycholate
TDCA: Taurodeoxycholate
THCA: Taurohyocholate
THDCA: Taurohyodeoxycholate
TLCA: Taurolithocholate
TUDCA: Tauroursodeoxycholate
UDCA: Ursodeoxycholate
wMCA: *omega*-Muricholate

## REFERENCES

1. Keum NN, Aune D, Greenwood DC, Ju W, Givannucci El. Calcium intake and cancer risk: Dose-response meta-analysis of prospective observational studies. Int J Cancer 2014;135:1940–1948.

2. Newmark HL, Yang K, Kurihara N, Fan K, Augenlicht LH, Lipkin M. Western-style diet-induced colonic tumors and their modulation by calcium and vitamin D in C57Bl/6 mice: a preclinical model for human sporadic colon cancer. Carcinogenesis. 2009;1:88–92.

3. Mariadason JM, Bordonaro M, Aslam F, Shi L, Kuraguchi M, Velcich A, et al. Downregulation of beta-catenin TCF signaling is linked to colonic epithelial cell differentiation. Cancer Res. 2001;61:3465–71.

4. Baron JA, Beach M, Mandel JS, van Stolk RU, Haile RW, Sandler RS, et al. Calcium supplements and colorectal adenomas. Polyp Prevention Study Group. Ann NY Acad Sci. 1999;889:138–45.

5. Grau MV, Baron JA, Sandler RS, Haile RW, Beach ML, Church TR, et al. Vitamin D, calcium supplementation, and colorectal adenomas: results of a randomized trial. J Natl Cancer Inst. 2003;95:1765–71.

6. Baron JA, Barry EL, Mott LA, Rees JR, Sandler RS, Snover DC, et al. A Trial of Calcium and Vitamin D for the Prevention of Colorectal Adenomas. N Engl J Med. 2015;373:1519–30.

7. Pommergaard HC, Burcharth J, Rosenberg J, Raskov H. Aspirin, Calcitriol, and Calcium Do Not Prevent Adenoma Recurrence in a Randomized Controlled Trial. Gastroenterology. 2016;150:114–122.

8. Crockett SD, Barry EL, Mott LA, Ahnen DJ, Robertson DJ, Anderson JC, et al. Calcium and vitamin D supplementation and increased risk of serrated polyps: results from a randomised clinical trial. Gut. 2018;pii: gutjnl-2017-315242.

9. Bolland MJ, Avenell A, Baron JA, Grey A, MacLennan GS, Gamble GD, et al. Effect of calcium supplements on risk of myocardial infarction and cardiovascular events: meta-analysis. BMJ. 2010;341:c3691.

10. Aslam MN, Paruchuri T, Bhagavathula N, Varani J. A mineral-rich red algae extract inhibits polyp formation and inflammation in the gastrointestinal tract of mice on a high-fat diet. Integr Cancer Ther. 20109:93-9.

11. Aslam MN, Bergin I, Naik M, Paruchuri T, Hampton A, Rehman M, et al. A multimineral natural product from red marine algae reduces colon polyp formation in C57BL/6 mice. Nutr Cancer. 2012;64:1020–8.

12. Aslam MN, Bhagavathula N, Paruchuri T, Hu X, Chakrabarty S, Varani J. Growth-inhibitory effects of a mineralized extract from the red marine algae, Lithothamnion calcareum, on Ca^2+^ -sensitive and Ca^2+^ -resistant human colon carcinoma cells. Cancer Lett. 2009;283:186–92.

13. Singh N, Aslam MN, Varani J, Chakrabarty S. Induction of calcium sensing receptor in human colon cancer cells by calcium, vitamin D and aquamin: Promotion of a more differentiated, less malignant and indolent phenotype. Mol Carcinog. 2015;54(7):543–53.

14. McClintock SD, Colacino JA, Attili D, Dame MK, Richter A, Reddy AR, et al. Calcium-Induced Differentiation of Human Colon Adenomas in Colonoid Culture: Calcium Alone versus Calcium with Additional Trace Elements. Cancer Prev Res (Phila). 2018;11:413–428.

15. Belkaid Y, Hand TW. Role of the microbiota in immunity and inflammation. Cell. 2014; 157:121–41.

16. Halfvarson J, Brislawn CJ, Lamendella R, Vázquez-Baeza Y, Walters WA, Bramer LM, et al. Dynamics of the human gut microbiome in inflammatory bowel disease. Nat Microbiol. 2017;2:17004.

17. Brennan CA, Garrett WS. Gut Microbiota, Inflammation, and Colorectal Cancer. Annu Rev Microbiol. 2016;70:395–411

18. Chaplin A, Parra P, Laraichi S, Serra F, Palou A. Calcium supplementation modulates gut microbiota in a prebiotic manner in dietary obese mice. Mol Nutr Food Res. 2016;60:468–80.

19. Aslam MN, Bassis CM, Zhang L, Zaidi S, Varani J, Bergin IL. Calcium Reduces Liver Injury in Mice on a High-Fat Diet: Alterations in Microbial and Bile Acid Profiles. PLoS One. 2016;11:e0166178.

20. Crowley EK, Long-Smith CM, Murphy A, Patterson E, Murphy K, O’Gorman DM, Stanton C, Nolan YM. Dietary Supplementation with a Magnesium-Rich Marine Mineral Blend Enhances the Diversity of Gastrointestinal Microbiota. Mar Drugs. 2018; 16; pii: E216.

21. Mello-Vieira J, Sousa T, Coutinho A, Fedorov A, Lucas SD, Moreira R, et al. Cytotoxic bile acids, but not cytoprotective species, inhibit the ordering effect of cholesterol in model membranes at physiologically active concentrations. Biochim Biophys Acta. 2013;1828:2152–63.

22. Bernstein H, Bernstein C, Payne CM, Dvorakova K, Garewal H. Bile acids as carcinogens in human gastrointestinal cancers. Mutat Res. 2005;589:47–65.

23. Wahlström A, Sayin SI, Marschall HU, Bäckhed F. Intestinal Crosstalk between Bile Acids and Microbiota and Its Impact on Host Metabolism. Cell Metab. 2016;24:41–50.

24. Ríos-Covián D, Ruas-Madiedo P, Margolles A, Gueimonde M, de Los Reyes-Gavilán CG, Salazar N. Intestinal Short Chain Fatty Acids and their Link with Diet and Human Health. Front Microbiol. 2016;7:185.

25. Diet History Questionnaire, Version 2.0. National Institutes of Health, Epidemiology and Genomics Research Program, National Cancer Institute. 2010. (Accessed June 21, 2019) https://epi.grants.cancer.gov/dhq2/

26. Dollive S, Chen YY, Grunberg S, Bittinger K, Hoffmann C, Vandivier L, et al. Fungi of the murine gut: episodic variation and proliferation during antibiotic treatment. PloS one. 2013;8(8):e71806.

27. Kozich JJ, Westcott SL, Baxter NT, Highlander SK, Schloss PD. Development of a dual-index sequencing strategy and curation pipeline for analyzing amplicon sequence data on the MiSeq Illumina sequencing platform. Appl Environ Microbiol. 2013; 79:5112–20.

28. Schloss PD, Westcott SL, Ryabin T, Hall JR, Hartmann M, Hollister EB, et al. Introducing mothur: open-source, platform-independent, community-supported software for describing and comparing microbial communities. Appl Environ Microbiol. 2009;75:7537–41.

29. Westcott SL, Schloss PD. OptiClust, an improved method for assigning amplicon-based sequence data to operational taxonomic units. MSphere. 2017;2(2):e00073–17.

30. Yue JC, Clayton MK. A similarity measure based on species proportions. Communications in Statistics-theory and Methods. 2005;34:2123–31

31. Segata N, Izard J, Waldron L, Gevers D, Miropolsky L, Garrett WS, Huttenhower C. Metagenomic biomarker discovery and explanation. Genome biology. 2011;12(6):R60.

32. Griffiths WJ, Sjövall J. Bile acids: analysis in biological fluids and tissues. J Lipid Res. 2010;51:23–41.

33. Demehri FR, Frykman PK, Cheng Z, Ruan C, Wester T, Nordenskjöld A, et al. HAEC Collaborative Research Group. Altered fecal short chain fatty acid composition in children with a history of Hirschsprung-associated enterocolitis. J Pediatr Surg. 2016;51:81–6.

34. Yang R, Chiang N, Oh SF, Serhan CN. Metabolomics-lipidomics of eicosanoids and docosanoids generated by phagocytes. Curr Protoc Immunol. 2011;Chapter 14:Unit 14.26.

35. Anderson MJ. A new method for non-parametric multivariate analysis of variance. Austral Ecology 2001; 26: 32–46

36. Eyssen HJ, De Pauw G, Van Eldere J. Formation of hyodeoxycholic acid from muricholic acid and hyocholic acid by an unidentified gram-positive rod termed HDCA 1 isolated from rat intestinal microflora. Appl Environ Microbiol. 1999;65:3158–63.

37. Louis P, Hold GL, Flint HJ. The gut microbiota, bacterial metabolites and colorectal cancer. Nat Rev Microbiol. 2014;12:661–72.

38. David LA, Maurice CF, Carmody RN, Gootenberg DB, Button JE, Wolfe BE, et al. Diet rapidly and reproducibly alters the human gut microbiome. Nature. 2014;505:559–63.

39. Sobhani I, Amiot A, Le Baleur Y, Levy M, Auriault ML, Van Nhieu JT, et al. Microbial dysbiosis and colon carcinogenesis: could colon cancer be considered a bacteria related disease? Therap Adv Gastroenterol. 2013;6:215–29.

40. Aslam MN, Bergin I, Naik M, Hampton A, Allen R, Kunkel SL, Rush H, Varani J. A multi-mineral natural product inhibits liver tumor formation in C57BL/6 mice. Biol Trace Elem Res. 2012;147(1-3):267–74.

41. Fujisaka S, Ussar S, Clish C, Devkota S, Dreyfuss JM, Sakaguchi M,et al. Antibiotic effects on gut microbiota and metabolism are host dependent. J Clin Invest. 2016;126(12):4430–4443.

42. Lemire JA, Harrison JJ, Turner RJ. Antimicrobial activity of metals: mechanisms, molecular targets and applications. Nat Rev Microbiol. 2013;11:371–84.

43. Haydel SE, Remenih CM, Williams LB. Broad-spectrum in vitro antibacterial activities of clay minerals against antibiotic-susceptible and antibiotic-resistant bacterial pathogens. J Antimicrob Chemother. 2008;61:353–61.

44. Morones-Ramirez JR, Winkler JA, Spina CS, Collins JJ. Silver enhances antibiotic activity against gram-negative bacteria. Sci Transl Med. 2013;5:190ra81.

45. Van der Meer R, Welberg JW, Kuipers F, Kleibeuker JH, Mulder NH, Termont DS, et al. Effects of supplemental dietary calcium on the intestinal association of calcium, phosphate, and bile acids. Gastroenterology. 1990;99:1653–9.

46. Eisenhofer R, Minich JJ, Marotz C, Cooper A, Knight R, Weyrich LS. Contamination in Low Microbial Biomass Microbiome Studies: Issues and Recommendations. Trends Microbiol. 2019;27:105–117.

47. Attili D, McClintock SD, Rizvi AH, Pandya S, Rehman H, Nadeem DM, et al. Calcium-induced differentiation in normal human colonoid cultures: Cell-cell / cell-matrix adhesion, barrier formation and tissue integrity. PLoS One. 2019;14:e0215122.

48. Orsi N. The antimicrobial activity of lactoferrin: current status and perspectives. Biometals. 2004;17:189–96.

49. Cellier MF, Courville P, Campion C. Nramp1 phagocyte intracellular metal withdrawal defense. Microbes Infect. 2007;9:1662–70.

50. Swaminath S, Um CY, Prizment AE, Lazovich D, Bostick RM. Combined Mineral Intakes and Risk of Colorectal Cancer in Postmenopausal Women. Cancer Epidemiol Biomarkers Prev. 2019;28:392–399.

